# Biological Pathways Linking Integrated Interventions to Infant Growth in the first 6 months of life: Findings from a Mediator Analysis of the WINGS Randomized Controlled Trial

**DOI:** 10.64898/2026.07.01.26357069

**Authors:** Ranadip Chowdhury, Devashish Tripathi, Sarita Devi, Sunita Taneja, Partha P Majumder, Tor A. Strand, R M Pandey, Anura V Kurpad, Souvik Mukherjee, Nita Bhandari

**Affiliations:** Society for Applied Studies, 45, Kalu Sarai, New Delhi, India; DBT and Wellcome India Alliance Clinical and Public Health Fellow, Hyderabad, India; Department of Physiology, St. John’s Medical College, Bengaluru, India; Human Genetics Unit, Indian Statistical Institute, Kolkata, West Bengal, India; Department of Research, Innlandet Hospital Trust, Lillehammer, Norway; Department of Biostatistics, Indian Council of Medical Research, New Delhi, India; Human Microbiome Research Laboratory, Biotechnology Research Innovation Council-National Institute of Biomedical Genomics, Kalyani, West Bengal, India; Regional Centre for Biotechnology, NCR Biotech Science Cluster, Faridabad, India

**Keywords:** Infant growth, SGA, integrated intervention package, mediation analysis, breastmilk, infant biomarkers, low- and middle-income countries

## Abstract

**Background:** Growth faltering during infancy remains a major public health concern in low- and middle-income countries. We examined whether birth size, infant gut microbiome, growth and inflammatory biomarkers, and breast milk macro and micronutrients mediated the effects of an integrated intervention package on infant growth at 6 months of life.

**Methods:** This mediation analysis was nested within the Women and Infants Integrated Interventions for Growth Study (WINGS), a factorial randomized controlled trial conducted in urban low- and middle-income communities in Delhi, India. The intervention package included nutrition, health, water, sanitation and hygiene (WASH), and psychosocial support delivered during the preconception, pregnancy, and early childhood. The analysis included 296 mother–infant dyads from the group received intervention from preconception throughout pregnancy and childhood and the group received routine care. Using a counterfactual causal mediation framework, we assessed the mediating roles of birth size, infant biomarkers at 3 months; insulin-like growth factor-1 [IGF-1], C-reactive protein [CRP], and *Bifidobacterium breve species*, and breast milk biomarkers at three months (vitamin B12 and total protein). Outcomes included length-for-age z-score (LAZ), weight-for-age z-score (WAZ), weight-for-length z-score (WLZ) at 6 months, and changes in these anthropometric indices between 3 and 6 months. Multiple mediators were evaluated simultaneously using parametric regression models adjusted for maternal height, maternal body mass index, family possess below poverty line card, and place of birth.

**Results:** The intervention package significantly improved WAZ at 6 months (total effect 0.31; 95% CI 0.04– 0.58). This effect was significantly mediated through birth size; reduction of small for gestational age (SGA) (average causal mediation effect 0.15; 95% CI 0.04–0.28), accounting for approximately 44% of the total effect.

**Conclusions:** Improved fetal growth, reflected by reduced SGA, was the principal biological pathway through which the integrated intervention package mediated infant growth at six months.

## Introduction

Growth faltering during early childhood remains a major public health concern in low- and middle-income countries, particularly in South Asia.(1) Growth failure during the first 2 years of age is associated with increased morbidity and mortality, impaired cognitive development, reduced educational attainment, and adverse health outcomes later in life.(2) Consequently, interventions delivered during the preconception, pregnancy, and early childhood periods have received considerable attention as strategies to improve child growth and development.(1)

However, the biological pathways through which such interventions affect growth remain incompletely understood. Infant growth is a complex process determined by multiple interrelated factors operating across the antenatal and postnatal periods, including fetal growth, gestational age at birth, nutritional status, immune function, gut microbiota composition, and the nutritional quality of breast milk.(3-8) Understanding whether these intermediate factors mediate the effects of interventions on subsequent growth can provide important insights into the mechanisms of action.

Birth size and gestational age at birth reflect the intrauterine environment and have been associated with later growth outcomes.(9) Early-life gut microbiota, particularly *Bifidobacterium* species, play a critical role in nutrient metabolism, immune maturation, and intestinal health and have been linked to infant growth.(10) Biomarkers such as insulin-like growth factor-1 (IGF-1), a key regulator of somatic growth, and C-reactive protein (CRP), a marker of systemic inflammation, represent biological pathways through which nutritional and environmental interventions affect growth.(11) Breast milk vitamin B12 is essential for cellular growth, neurological development, and hematopoiesis, while breast milk protein contributes directly to infant nutritional status and growth.(12, 13) Despite the biological plausibility of these pathways, evidence regarding their mediating role in intervention effects into improved early childhood growth outcome is limited. Traditional mediation analyses often evaluate individual mediators separately, potentially overlooking the complex interrelationships among multiple biological pathways. Recent advances in causal inference methods enable the simultaneous assessment of multiple mediators within a counterfactual framework, allowing estimation of both overall indirect effects and mediator-specific contributions. (14)

The present study, nested within the WINGS trial,(15) aimed to investigate the mechanisms linking an integrated intervention package delivered during the preconception, pregnancy, and early childhood with infant growth outcomes at six months of age. We evaluated whether birth size, infant biomarkers at 3 months (CRP, IGF-1, relative abundance of Bifidobacterium breve in infant gut), and breast milk biomarkers at 3 months (vitamin B12 and total protein) mediated the intervention effect on growth outcomes, including length-for-age z-score (LAZ), weight-for-age z-score (WAZ), weight-for-length z-score (WLZ), and changes in these anthropometric indicators between 3 and 6 months.

## Methodology

### Ethics statement

Ethical clearance for this study was granted by the Ethics Review Committee of the Society for Applied Studies in New Delhi, India (Ref no. SAS/ERC/WINGS-Sub Study/2020). The study complied with ethical standard outlined in Declaration of Helsinki. This study was registered with the Clinical Trial Registry of India under the identifier CTRI/2020/10/028770. Written informed consent was secured primarily from mothers before their inclusion in the study.

### Primary study design

This was a factorial randomized controlled trial (RCT) conducted in low- and middle-income neighbourhoods of Delhi, India. Eligible women of reproductive age (18–30 years), identified through door-to-door survey were enrolled and randomized to receive either the preconception intervention package or routine care. They were followed for 18 months. If they were identified as pregnant in this period (ultrasonographic confirmation of pregnancy), they were re-randomized either to receive the pregnancy and early childhood intervention package or routine care. All pregnant women were followed until delivery, and their children until they reached 24 months of age. This two-step randomization resulted in four groups: preconception and pregnancy interventions (A), preconception interventions only (B), pregnancy and early childhood interventions only (C), and no preconception interventions, with routine pregnancy and early childhood care (D). The interventions, under the four domains of health, nutrition, WASH, and psychosocial care and support, are briefly described below.

In the pre-conception period, women were screened and treated for medical conditions, including anemia. All received one tablet of iron and folic acid (IFA) daily and weekly multiple micronutrients (MMN) supplements to meet ½ to ¾ of the recommended daily allowance (RDA). They also received an albendazole tablet twice a year. All women were counseled on adequate diets, positive thinking, problem-solving skills, and menstrual hygiene.

In the pregnancy and early childhood interventions, women received monthly antenatal care, screening, and treatment for medical conditions, including anemia. Women received daily IFA, MMN (∼1 RDA), calcium, and vitamin D throughout pregnancy. Albendazole was given once during pregnancy. Women with BMI < 25 kg/m2 in the second and third trimesters received food supplements. Women with inadequate weight gain (IWG) were identified based on the Institute of Medicine guidelines for gestational weight gain (GWG), and nutritional counseling and extra food supplements were provided.

Water filters, hand washing stations, soap, and disinfectants were provided to all families, in addition to counselling on sanitation and hygiene. Mothers received counselling on exclusive breastfeeding during the first 6 months of the infant’s age. Additional visits were made for babies born preterm, LBW, and for mothers with breastfeeding problems. Nutritional supplementation included vitamin D for all infants, iron for very low and low birth weight infants, and daily snacks and supplements for mothers. (15)

### Participants of the current study

Between 1st December 2020 and 30th December 2021, 296 mother-infant dyads were selected consecutively when the infants completed 6 months of age from Group A and D as defined in the primary WINGS study. An independent statistician provided a list to ensure unbiased group allocation. All outcome assessments were conducted by a separate team that was not involved in the intervention delivery and was blinded to group assignments before the measurement process. Infant gut microbiome was analysed by high-throughput sequencing of the V3-V4 region of 16S rRNA gene. (16) Serum IGF-1 was analyzed by electrochemiluminescence immunoassay on the Cobas e 601 analyzer (Roche Diagnostics, Mannheim, Germany) using the Elecsys IGF-1 kit (Roche Diagnostics, Ref No. 07475896190), and Serum CRP was measured by immunoturbidometric based assay on the Cobas c 501 analyzer (Roche Diagnostics, Mannheim, Germany).(17) Total protein concentrations in human breast milk samples were quantified using the bicinchoninic acid (BCA) assay (Pierce™ BCA Protein Assay Kit, Cat# 23225, Thermo Scientific, USA), following the manufacturer’s protocol. Vitamin B12 (holotranscobalamin) concentrations were quantified by chemiluminescence immunoassay using Cobas e411 (Roche Diagnostics) after defatting the breastmilk samples.

### Statistical analysis

We performed causal mediation analysis to decompose the total effect of intervention package on the infant growth outcomes into natural direct and natural indirect effect. We considered the intervention package provided to mother and infants as exposure. We included mediators infant birth size (preterm or term), birth size (SGA or appropriate for gestational age (AGA); infant biomarkers at 3 months, including relative abundance of Bifidobacterium breve in infant gut, CRP, and IGF-1; and breast milk macro- and micronutrients at 3 months, total protein and vitamin B12. The infant growth outcomes were LAZ, WAZ, and WLZ at 6 months, as well as the changes in LAZ, WAZ, and WLZ between 3 and 6 months. We considered the maternal height and BMI, family possess bpl card, and place of birth as the confounders for exposure-outcome, exposure-mediator, and mediator-outcome relationship. Mediation analysis was conducted under the counterfactual framework, assuming sequential ignorability and using parametric regression models. (18) As illustrated in the DAG (Figure 1), we used the R package *multimediate* to incorporate multiple mediators simultaneously within the causal mediation analysis framework. This approach allowed us to jointly estimate the overall indirect effect through all mediators and to quantify the mediator-specific indirect effects. Multiple linear regression models were used for all outcomes and mediators, except birth size, for which a probit regression model was applied. All mediator and outcome models were adjusted for the specified confounders. Intervention–mediator interaction terms were not included in the outcome models, as the estimated interaction effects were largely non-significant in preliminary analyses. We performed 1000 Monte Carlo draws for a quasi-Bayesian approximation to estimate the total effect, average direct effect, average indirect effect, and the proportion of the total effect explained by mediation, along with 95% confidence intervals and p-values.

**Figure 1:**
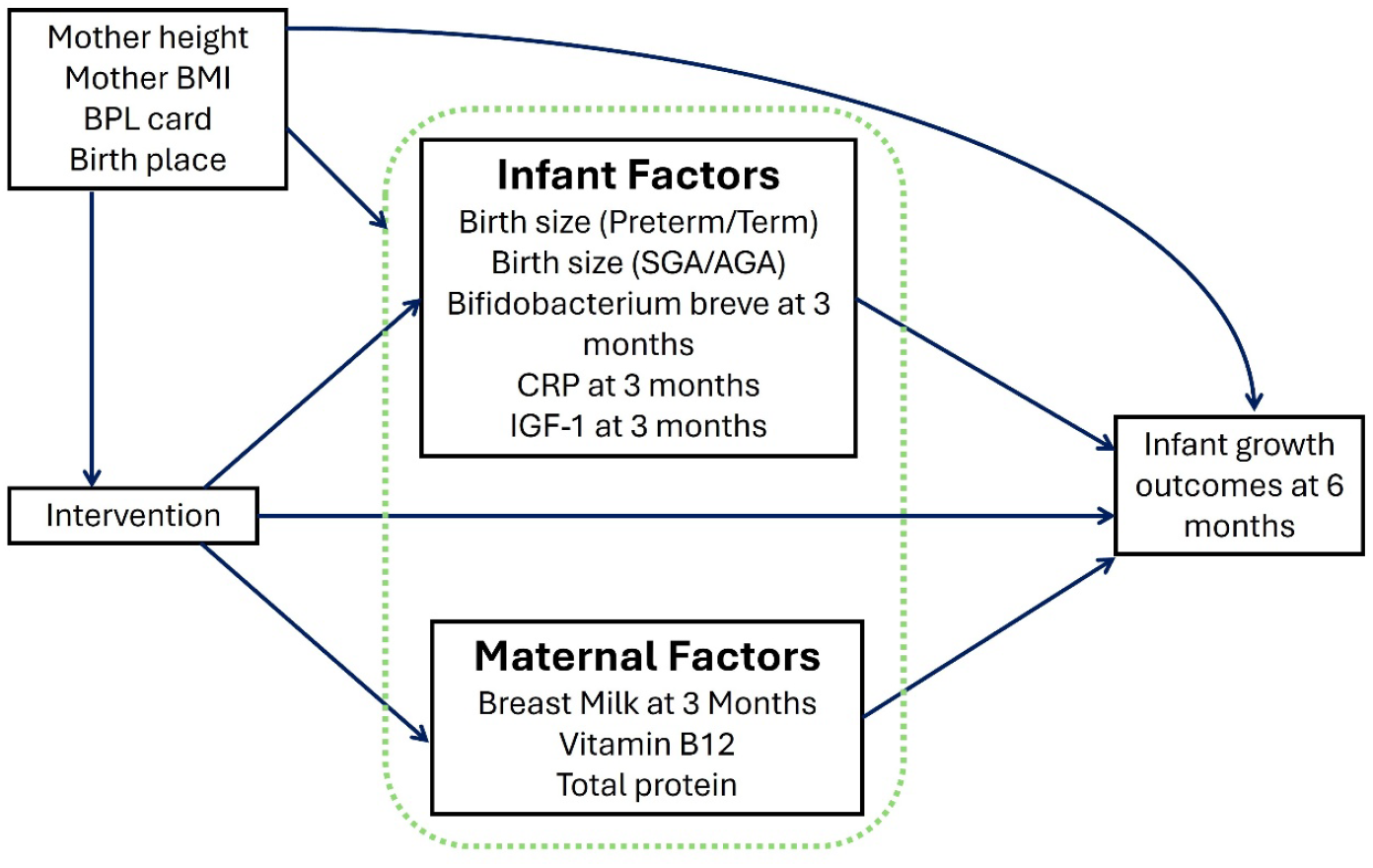
Causal pathway diagram.

## Results

Between December 2020 and December 2021, 296 infants (141 in the preconception, pregnancy, and early childhood intervention group, and 155 in the routine care group). The baseline characteristics of the women were similar, except for the proportion of underweight women, the number of families possessing a below the poverty line (BPL) card, and the place of birth (Table 1).

**Table 1:** Baseline characteristics of women at recruitment.

|  | Preconception, pregnancy and<br>early childhood (Group A)(N =<br>141) | Routine care<br>(Group D)(N =<br>155) |
| --- | --- | --- |
| Women age (y); Mean (SD) | 23.9 (3.1) | 24.2 (3.0) |
| Women height (cm); Mean (SD) | 151.7 (5.5) | 152.5 (5.5) |
| Women height <150 cm | 60 (42.6) | 47 (30.3) |
| BMI category (kg/m<sup>2</sup>) |  |  |
| ≥25 | 34 (24.1) | 36 (23.2) |
| 18.5–24.99 | 87 (61.7) | 92 (59.4) |
| <18.5 | 20 (14.2) | 27 (17.4) |
| Joint or extended family | 86 (61.0) | 105 (67.7) |
| Women schooling ≥12 y | 81 (57.4) | 85 (54.8) |
| Homemaker | 137 (97.2) | 148 (95.5) |
| Family has below poverty line card | 5 (3.5) | 10 (6.5) |
| Family covered by health insurance<br>scheme | 7 (5.0) | 15 (9.7) |
| Place of birth |  |  |
| Large hospital | 115 (81.6) | 75 (48.4) |
| Small hospital or birthing center | 23 (16.3) | 66 (42.6) |
| Home birth | 3 (2.1) | 14 (9.0) |

Data presented as Mean(SD) or count (%) as appropriate

**Table 2** presents the direct and indirect effect of intervention package on infant growth outcomes at 6 months. The total effect of intervention package.

**Table 2:**
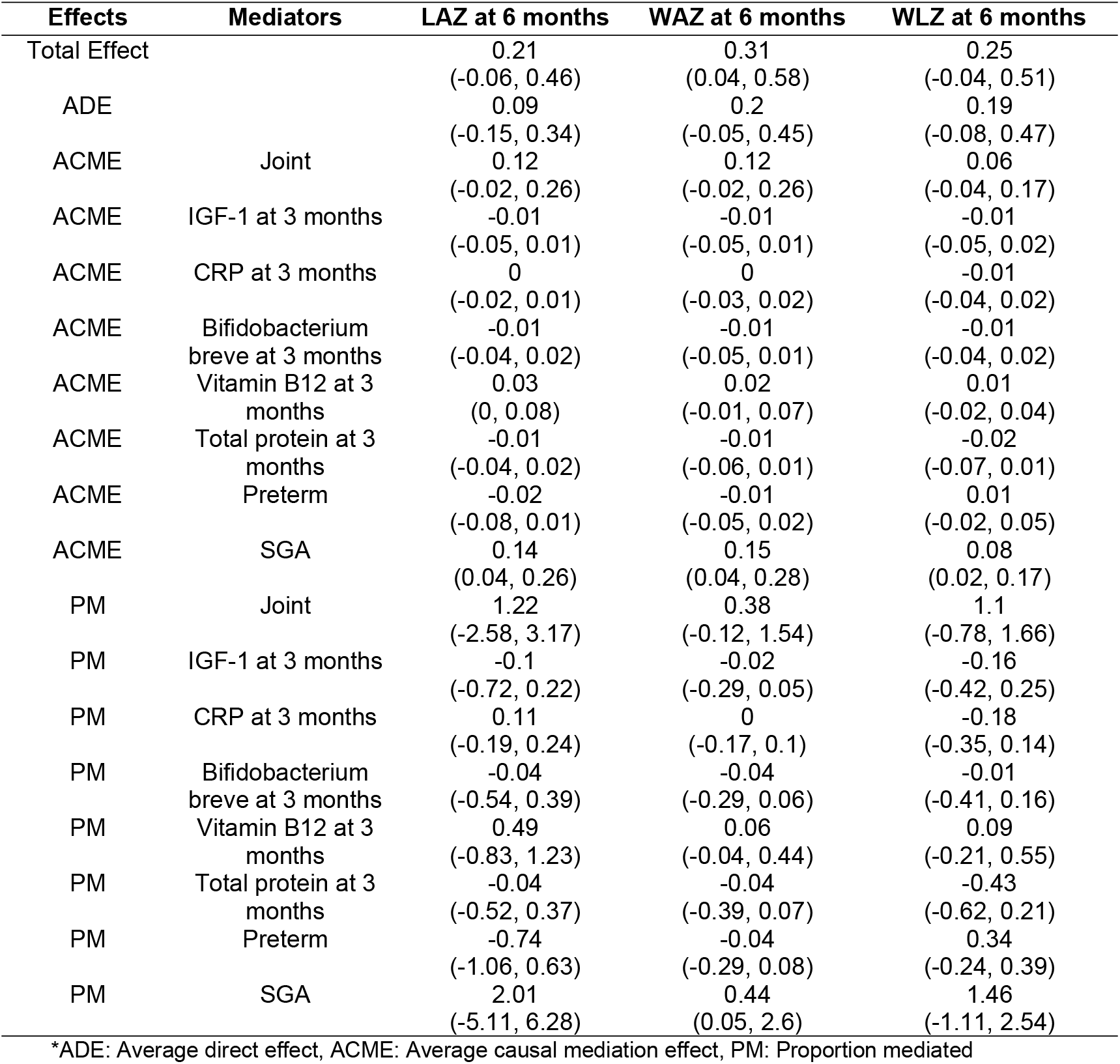
Mediation analysis for infant growth outcomes at 6 months (N=296)

| Effects | Mediators | LAZ at 6 months | WAZ at 6 months | WLZ at 6 months |
| --- | --- | --- | --- | --- |
| Total Effect |  | 0.21<br>(-0.06, 0.46) | 0.31<br>(0.04, 0.58) | 0.25<br>(-0.04, 0.51) |
| ADE |  | 0.09<br>(-0.15, 0.34) | 0.2<br>(-0.05, 0.45) | 0.19<br>(-0.08, 0.47) |
| ACME | Joint | 0.12<br>(-0.02, 0.26) | 0.12<br>(-0.02, 0.26) | 0.06<br>(-0.04, 0.17) |
| ACME | IGF-1 at 3 months | -0.01<br>(-0.05, 0.01) | -0.01<br>(-0.05, 0.01) | -0.01<br>(-0.05, 0.02) |
| ACME | CRP at 3 months | 0<br>(-0.02, 0.01) | 0<br>(-0.03, 0.02) | -0.01<br>(-0.04, 0.02) |
| ACME | Bifidobacterium breve at 3 months | -0.01<br>(-0.04, 0.02) | -0.01<br>(-0.05, 0.01) | -0.01<br>(-0.04, 0.02) |
| ACME | Vitamin B12 at 3 months | 0.03<br>(0, 0.08) | 0.02<br>(-0.01, 0.07) | 0.01<br>(-0.02, 0.04) |
| ACME | Total protein at 3 months | -0.01<br>(-0.04, 0.02) | -0.01<br>(-0.06, 0.01) | -0.02<br>(-0.07, 0.01) |
| ACME | Preterm | -0.02<br>(-0.08, 0.01) | -0.01<br>(-0.05, 0.02) | 0.01<br>(-0.02, 0.05) |
| ACME | SGA | 0.14<br>(0.04, 0.26) | 0.15<br>(0.04, 0.28) | 0.08<br>(0.02, 0.17) |
| PM | Joint | 1.22<br>(-2.58, 3.17) | 0.38<br>(-0.12, 1.54) | 1.1<br>(-0.78, 1.66) |
| PM | IGF-1 at 3 months | -0.1<br>(-0.72, 0.22) | -0.02<br>(-0.29, 0.05) | -0.16<br>(-0.42, 0.25) |
| PM | CRP at 3 months | 0.11<br>(-0.19, 0.24) | 0<br>(-0.17, 0.1) | -0.18<br>(-0.35, 0.14) |
| PM | Bifidobacterium breve at 3 months | -0.04<br>(-0.54, 0.39) | -0.04<br>(-0.29, 0.06) | -0.01<br>(-0.41, 0.16) |
| PM | Vitamin B12 at 3 months | 0.49<br>(-0.83, 1.23) | 0.06<br>(-0.04, 0.44) | 0.09<br>(-0.21, 0.55) |
| PM | Total protein at 3 months | -0.04<br>(-0.52, 0.37) | -0.04<br>(-0.39, 0.07) | -0.43<br>(-0.62, 0.21) |
| PM | Preterm | -0.74<br>(-1.06, 0.63) | -0.04<br>(-0.29, 0.08) | 0.34<br>(-0.24, 0.39) |
| PM | SGA | 2.01<br>(-5.11, 6.28) | 0.44<br>(0.05, 2.6) | 1.46<br>(-1.11, 2.54) |

\*ADE: Average direct effect, ACME: Average causal mediation effect, PM: Proportion mediated

**WAZ:** The intervention package had a statistically significant positive total effect on WAZ (β = 0.31, 95% CI: 0.04–0.58). Mediation analysis demonstrated a significant indirect effect through SGA status (β = 0.15, 95% CI: 0.04–0.28), indicating that improvements in fetal growth contributed substantially to the intervention effect. Approximately 44% of the total effect on WAZ was mediated through SGA.

**WLZ:** The intervention package showed a positive, although non-significant, total effect on WLZ (β = 0.25, 95% CI: −0.04 to 0.51). However, mediation analysis identified a significant indirect effect through SGA (β = 0.08, 95% CI: 0.02–0.17), suggesting that the intervention influenced WLZ primarily by reducing the risk of SGA at birth.

**LAZ:** No statistically significant total effect, direct effect, or overall joint indirect effect of the intervention package on LAZ was observed. Nevertheless, among the individual mediators evaluated, SGA demonstrated a significant positive average causal mediation effect (ACME) on LAZ (β = 0.14, 95% CI: 0.04–0.26), indicating that improvements in birth size partially mediated the relationship between the intervention package and subsequent linear growth, despite the absence of a significant overall intervention effect.

**Table 3** presents the mediation analyses for change in LAZ, WAZ, and WLZ scores between 3 and 6 months. No statistically significant total or average direct effects were observed for any of the three changes in outcomes. The joint indirect effects were also not significant. None of the mediators showed evidence of significant mediation, and all estimated proportions mediated were statistically non-significant.

**Table 3:** Mediation analysis for difference of infant growth outcomes at 6 months and 3 months (N = 296)

| Effects | Mediators | LAZ difference<br>between 6 to 3<br>months | WAZ difference<br>between 6 to 3<br>months | WLZ difference<br>between 6 to 3<br>months |
| --- | --- | --- | --- | --- |
| Total Effect |  | -0.03<br>(-0.18, 0.13) | 0.01<br>(-0.12, 0.14) | 0.1<br>(-0.08, 0.29) |
| ADE |  | -0.05<br>(-0.2, 0.1) | 0.02<br>(-0.11, 0.15) | 0.11<br>(-0.08, 0.29) |
| ACME | Joint | 0.02<br>(-0.04, 0.09) | -0.01<br>(-0.06, 0.05) | -0.01<br>(-0.07, 0.06) |
| ACME | IGF-1 at 3<br>months | -0.01<br>(-0.03, 0.01) | 0<br>(-0.02, 0.01) | 0<br>(-0.02, 0.02) |
| ACME | CRP at 3<br>months | 0<br>(-0.01, 0.01) | 0<br>(-0.01, 0.01) | 0<br>(-0.02, 0.01) |
| ACME | Bifidobacterium<br>breve at 3<br>months | 0<br>(-0.01, 0.02) | 0<br>(-0.02, 0.01) | -0.01<br>(-0.04, 0.01) |
| ACME | Vitamin B12 at<br>3 months | -0.01<br>(-0.03, 0.01) | 0<br>(-0.01, 0.02) | 0.02<br>(0, 0.05) |
| ACME | Total protein at<br>3 months | 0<br>(-0.02, 0.02) | 0.01<br>(0, 0.03) | 0.01<br>(-0.01, 0.04) |
| ACME | Preterm | 0.03<br>(-0.01, 0.09) | 0.02<br>(-0.01, 0.06) | -0.02<br>(-0.05, 0.01) |
| ACME | SGA | 0<br>(-0.03, 0.02) | -0.03<br>(-0.07, -0.01) | -0.01<br>(-0.04, 0.02) |
| PM | Joint | -2.37<br>(-3.99, 5.99) | 0.63<br>(-5.97, 5.06) | 0.26<br>(-2.45, 3.23) |
| PM | IGF-1 at 3<br>months | 0.2<br>(-1.43, 1.13) | 0.2<br>(-1.01, 1.37) | -0.11<br>(-0.73, 0.75) |
| PM | CRP at 3<br>months | 0.2<br>(-0.6, 0.41) | 0.22<br>(-0.6, 0.59) | -0.01<br>(-0.37, 0.38) |
| PM | Bifidobacterium<br>breve at 3<br>months | -0.24<br>(-0.94, 0.94) | 0.35<br>(-1.56, 1.02) | -0.11<br>(-1.44, 1.12) |
| PM | Vitamin B12 at<br>3 months | -0.01<br>(-1.27, 1.04) | -0.04<br>(-1.27, 1.18) | -0.02<br>(-0.97, 2.05) |
| PM | Total protein at<br>3 months | -0.45<br>(-1, 1.03) | -0.45<br>(-2.87, 2.08) | 0.33<br>(-0.94, 1.71) |
| PM | Preterm | -0.35<br>(-4.92, 5.98) | -0.54<br>(-6.1, 4.54) | -0.1<br>(-1.83, 1.07) |
| PM | SGA | 0.1<br>(-1.4, 1.8) | 0.7<br>(-5.54, 9.83) | -0.23<br>(-1.53, 1.22) |

\*ADE: Average direct effect, ACME: Average causal mediation effect, PM: Proportion mediated

## Discussion

This study assessed the biological pathways through which an integrated intervention package delivered during the preconception, pregnancy, and early childhood mediated infant growth outcomes at six months of age. Using a counterfactual causal mediation framework, we found that improved fetal growth, reflected by a reduction in SGA, was the principal pathway linking the integrated intervention to improved infant growth. SGA significantly mediated the intervention effect on WAZ, accounting for approximately 44% of the total effect. Preterm birth, infant growth and inflammatory biomarkers (IGF-1 and CRP), infant gut microbiota (Bifidobacterium breve), and breast milk vitamin B12 and total protein measured at 3 months of life did not significantly mediate intervention effects.

Our finding that SGA was the primary mediator of the intervention effect is biologically plausible because birth size integrates the cumulative effects of the intrauterine nutritional, metabolic, endocrine, placental, and inflammatory environment. Maternal nutritional status before and during pregnancy influences placental development, vascularization, nutrient transport, and endocrine signaling which are critical determinants of fetal growth and birth size.(19-21) The integrated intervention package in the WINGS trial was initiated from preconception and continued throughout pregnancy, providing nutritional supplementation, micronutrients, infection prevention and management, enhanced antenatal care, and psychosocial support during a critical developmental window when placental function and fetal nutrient transfer are established. The primary WINGS trial demonstrated a 31% reduction in SGA among participants who received the intervention package from preconception through pregnancy compared with those receiving routine care.(15) Our mediation analysis findings therefore suggest that the beneficial effects of the integrated intervention on WAZ during early infancy were established predominantly through improvements in fetal growth rather than through the postnatal biological pathways measured at three months. The absence of significant mediation through postnatal biomarkers, including IGF-1, CRP, Bifidobacterium breve, breast milk vitamin B12, and total protein, may indicate that the major growth benefit of the intervention was established before birth rather than through the postnatal biological pathways measured at 3 months of life.

The strengths of this study include the use of a counterfactual mediation framework, simultaneous evaluation of multiple biologically plausible mediators, and adjustment for important maternal and birth-related confounders. The analysis provides a more comprehensive assessment of potential pathways than traditional single-mediator approaches. Furthermore, the study examined both attained growth outcomes and growth changes, offering insights into different dimensions of infant growth.

Several limitations should be considered when interpreting the findings. First, mediation analyses rely on the assumption of no unmeasured confounding between exposure, mediators, and outcomes, an assumption that cannot be fully verified. Second, the relatively modest sample size may have limited the ability to detect small indirect effects, particularly for individual mediators.

Third, mediators were measured at a single time point, which may not adequately capture biological processes that vary over time. Finally, the selected mediators do not encompass the full range of mechanisms through which maternal and child interventions may influence growth. In conclusion, this mediation analysis of the WINGS randomized controlled trial demonstrated that improved fetal growth, reflected by a reduction in SGA, is the primary biological pathway through which an integrated intervention package delivered from preconception through pregnancy and early childhood improves infant WAZ at six months of age. Approximately 44% of the intervention effect on WAZ was mediated through SGA, highlighting the critical importance of optimizing fetal growth during the prenatal period. Infant inflammatory and growth biomarkers, gut microbiota, and breast milk vitamin B12 and total protein measured at three months did not explain the observed intervention effects. These findings suggest that the benefits of integrated maternal and child health interventions may be established before birth through improvements in the intrauterine environment rather than through the postnatal biological pathways examined in this study.

## Data Availability

Individual requests will be considered on a case-by-case basis. The request for data should be accompanied by a detailed proposal describing the scientific questions to be addressed. Proposals should be submitted to RC. Dataset used in this study for analysis can be accessed using https://figshare.com/s/109de02b7afafc0fb3b5

https://doi.org/10.6084/m9.figshare.32820323

## Data Sharing

Individual requests will be considered on a case-by-case basis. The request for data should be accompanied by a detailed proposal describing the scientific questions to be addressed.

Proposals should be submitted to RC.

## Author Contributions

**Conceptualization:** Ranadip Chowdhury

**Data curation:** Ranadip Chowdhury

**Formal analysis:** Devashish Tripathi

**Methodology:** Ranadip Chowdhury

**Supervision:** Ranadip Chowdhury

**Writing – original draft:** Ranadip Chowdhury, Devashish Tripathi

**Technical guidance:** Partha P Majumder, Anura V Kurpad, Tor A. Strand, Ravindra Mohan Pandey, Sunita Taneja, Sarita Devi, Souvik Mukherjee and Nita Bhandari

**Writing – review & editing:** All authors

## Acknowledgement

We deeply acknowledge the contribution of the participants and their families and are thankful to the community leaders for their cooperation.

